# SafeSwab, a sample collection and dispensing device for near-patient testing

**DOI:** 10.1101/2021.08.20.21262386

**Authors:** Thomas E. Grys, Kathrine McAulay, Darrell Ingram, Craig Duffy, Ashley Williams, Siddarth Arumugam, Jiawei Ma, Uzay Macar, Guangxing Han, Andy Reynolds, Shih-Fu Chang, Samuel K. Sia, Ken Mayer

## Abstract

**Background:** The COVID-19 pandemic has accelerated the pace of innovation around virtual care visits and testing technology. Here we present the SafeSwab (Safe Health Systems, Los Angeles, CA), an integrated, universal sample collection and dispensing device that is designed to minimize user error and enable rapid testing in a point of care or self-testing format.

**Methods:** The SafeSwab was used with the Safe Health Systems HealthCheck digital health application to enable self-testing by patients using lateral flow tests for SARS-CoV-2 antigen or for antibodies against SARS-CoV-2.

**Results:** Patients (n=74) using the SafeSwab produced a valid rapid test result in 96% of attempts, and 96% of patients felt confident that they had collected a good sample. The Safe HealthCheck app has an integrated image analysis algorithm, AutoAdapt LFA, that interprets a picture of a rapid test result, and the algorithm interpreted the result correctly 100% of the time.

**Conclusion:** The SafeSwab was found to be versatile and easy to use for both self-collected nasal sampling as well as fingerstick blood sampling. The use of Safe Health Systems HealthCheck app allows an integrated solution for patient instruction and test interpretation

## Introduction

The SARS-CoV-2 pandemic has spurred an intense period of research and innovation like none ever seen before. The U.S. Food and Drug Administration (FDA) has recently authorized antigen tests for use in non-laboratory settings, including the home (1). Although these tests are relatively simple to perform for those who have laboratory experience or medical training, they are dependent on adequate specimen collection and adequate performance of several time and volume-dependent steps. The motivation to approve home-based testing was the urgency of the pandemic. It is likely, however, that tests for other infectious diseases and chronic diseases will be cleared through the 510(k) and premarket approval (PMA) process for use in non-laboratory settings. The most successful among them will be those that optimize the testing process in a way that increases the robustness of the test and minimizes the chance of user error.

We introduce here the SafeSwab, an integrated sample collection and dispensing device that is broadly applicable to near-patient testing, whether point-of-care or home use. The device is designed to minimize errors of collection, handling, and measuring. The SafeSwab was used in combination with a digital health solution called Safe HealthCheck (Safe Health Systems), to allow patients to test themselves. Safe HealthCheck is a smart phone application that integrates symptom checking, virtual consults, animated instructional videos, and an automated machine learning algorithm (AutoAdapt LFA) for automated test interpretation (Arumugam et al., under review). Here we outline a usability study in the context of COVID-19, where our primary outcome was participant satisfaction with the devise and test validity. Specific assay performance is not addressed here.

## Methods

### SafeSwab

The SafeSwab (Figure 1) is a collection device that allows for integrated sample collection and dispensing. After using a standard lancet, the absorbent tip of the SafeSwab can be used to collect a fingerstick blood sample. Or, the swab tip can be extended to reveal approximately 1 cm of swab surface to collect a nasal sample. Distal to the tip is a reservoir, which can be filled with any buffer to suit the test being performed. Twisting the reservoir releases the buffer to flow down the barrel, carrying the sample out of the absorbent tip. When held over a rapid testing device, the sample can be placed directly into the sample inlet.

**Figure 1.**
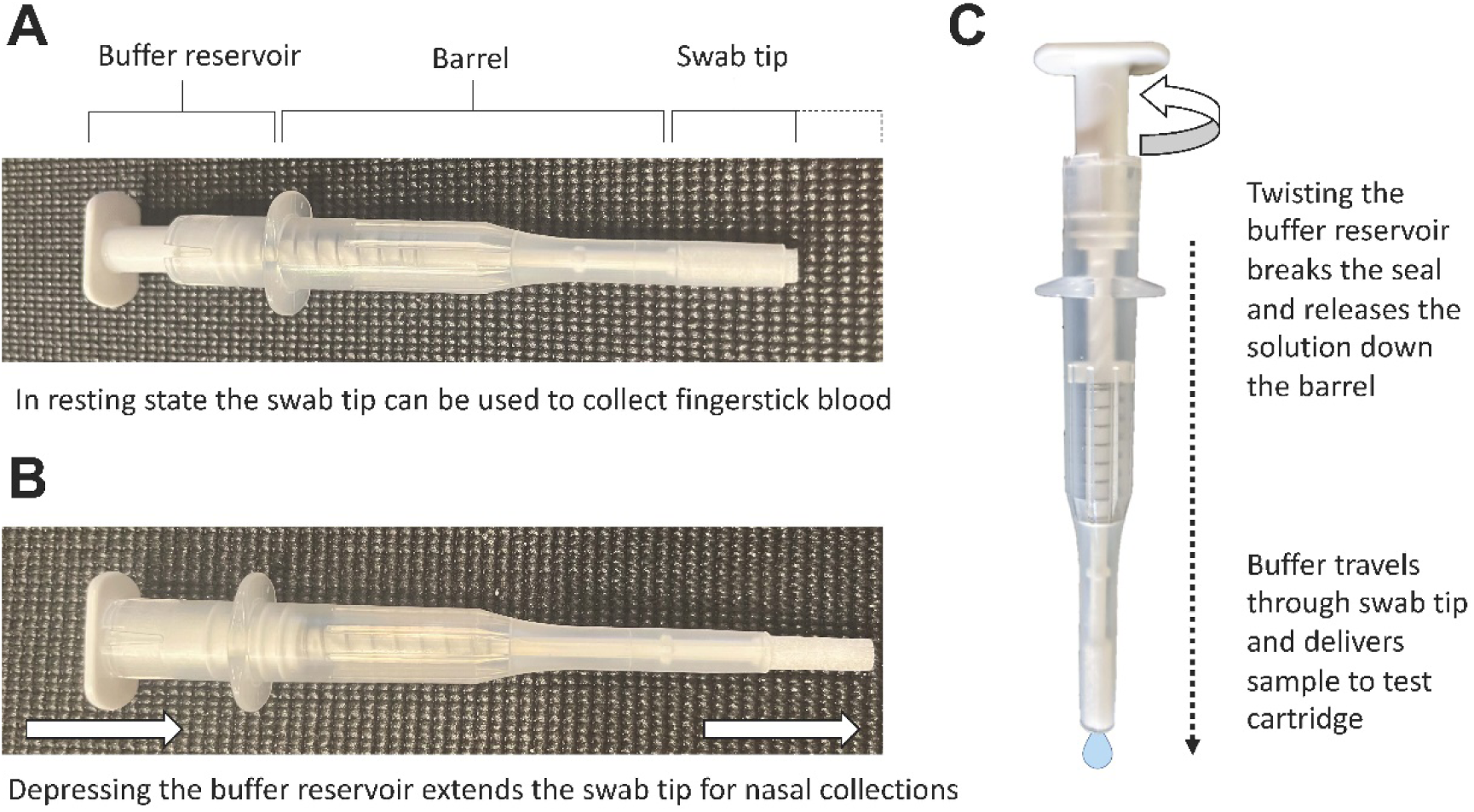
SafeSwab in resting state for fingerstick blood sampling (A), where blood is absorbed directly onto the swab tip, and with the swab extended for nasal sampling (B). Twisting the buffer reservoir (C), distal to the swab tip, breaks the seal, releasing buffer down the shaft and through the swab tip, carrying the sample into the testing kit.

### Approach

To evaluate the usability of the SafeSwab, we recruited individuals undergoing standard of care (SOC) SARS-CoV-2 testing [nasopharyngeal (NP) swab tested using one of two Emergency Use Authorized (EUA) PCR methods, RealTi*m*e SARS-CoV-2 assay on the *m*2000 (Abbott Laboratories) system or Alinity m SARS-CoV-2 assay (Abbott)]

### Antigen testing population

Participants in the antigen testing arm were recruited via the drive-through COVID-19 testing site at an academic hospital campus. After providing informed consent (IRB protocol 20-010688), participants were asked to remain in their vehicle (to simulate a home environment) and were provided with a small tray containing a Flowflex™ SARS-CoV-2 Antigen Rapid Test cartridge (ACON Laboratories), a SafeSwab pre-filled with Flowflex™ SARS-CoV-2 Antigen Rapid Test buffer (ACON), and a mobile phone with the Safe Health Systems HealthCheck application installed.

### Antibody testing population

Participants in the antibody testing arm were contacted ≥ 2 weeks after a positive PCR result and invited to return ≥ 3 weeks from their positive PCR result and ≥ 2 weeks from symptom resolution. After informed consent (IRB protocol 20-004544), participants were asked to remain in their vehicle (to simulate a home environment) and were provided with a small tray containing an ACON SARS-CoV-2 IgG/IgM Rapid Test cartridge (ACON), a SafeSwab pre-filled with ACON SARS-CoV-2 IgG/IgM Rapid Test buffer, alcohol prep pad, lancet, bandage, and mobile phone.

### Self-testing

Participants in both studies used the Safe HealthCheck phone application to complete the testing process. First the app prompts the user to scan a QR code on the test cartridge pouch, initiating the cognate animated instructional video (motion504, Minneapolis, MN). The video shows the user the entire testing process. After the video, still images from the video instructions with accompanying text take the user through the testing process step by step on subsequent screens (Figure 2). The app includes a built-in timer and subsequent prompt for the user to take a picture of the cartridge (also via the app). The test image is sent to an Amazon Web Server where it is processed by the AutoAdapt LFA image analysis algorithm, which uses machine learning to detect the test strip and any bands present, then uses a lookup table containing information on positions of test vs. control lines to interpret the result (2). During the study, the result was not returned to the user, but was stored in a de-identified database for documentation purposes.

**Figure 2.**
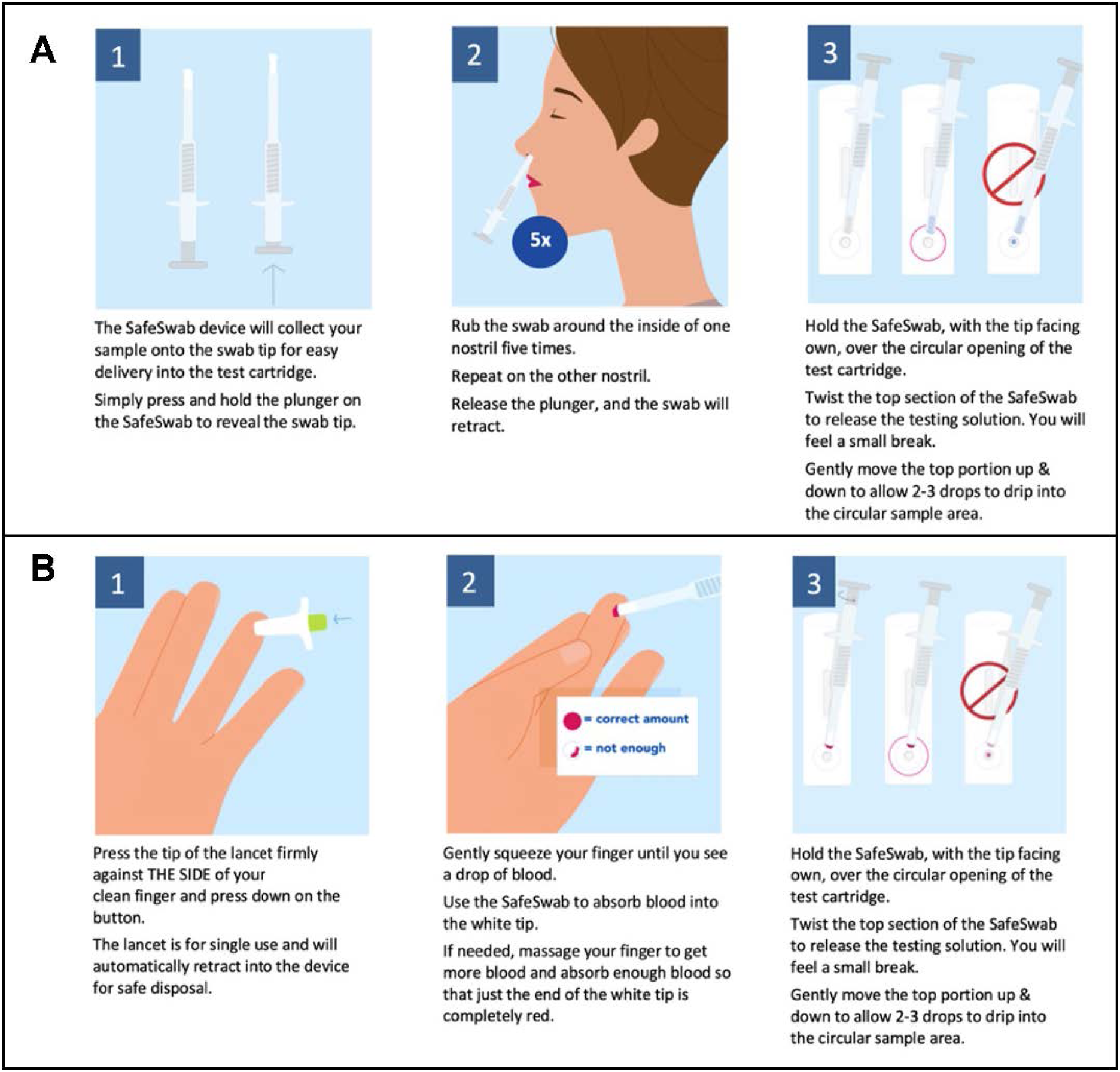
Representative images from the instructional sequences provided to users in this study for antigen (A) and antibody (B) testing.

### Survey

Each participant was asked to complete a usability study at the end of their experience. During the survey, participants were asked to provide their age and maximum educational level and to answer 5-7 usability questions anonymously.

### Analysis

The survey results were collated to assess SafeSwab Usability. To assess SafeSwab performance, as opposed to specific assay performance, the primary metric assessed was result validity, i.e., tests generating a control line were considered valid, while those failing to generate a control line were considered invalid (See Supplementary Figure 1 for representative images). Additionally, dilution series were performed using discarded nasopharyngeal swab samples in 3 mL viral transport medium (VTM) from the hospital Microbiology laboratory. Two samples from patients with acute disease were diluted 1:10 in a series into sterile saline. For each dilution, 10 μL and 30 μL was transferred to a microtiter plate. The SafeSwab was used to absorb the sample, and then the buffer chamber activated to dispense 2-3 drops into the sample well on the antigen kits

## Results and Discussion

Of the 40 participants in the antigen testing arm, 37 produced a valid result and 3 produced an invalid result (control line missing). Among the 37 valid results, 34 matched the PCR results (32 true negatives and 2 true positives), while 3 produced unexpected results (2 false negatives and one false positive). The AutoAdapt LFA image interpretation algorithm correctly called all the tests in agreement with human observation, including the three invalid results. True positives demonstrated crossing thresholds (Ct) of 27.85 (Alinity m SARS-CoV-2 assay) and 5.37 [RealTi*m*e SARS-CoV-2 assay, which produces results approximately 14 Ct lower than Alinity m SARS-CoV-2 assay (3)]. False negatives demonstrated Ct values of 26.18 and 40.65 (Alinity m SARS-CoV-2 assay). This data is consistent with accounts of reduced sensitivity in LFA antigen assays when compared with PCR (4-7). Only two PCR positive patients were captured in the antigen testing arm of this study. A random sampling approach was employed, where both symptomatic and non-symptomatic patients were invited to participate, although it is possible that uptake may have been lower among those feeling unwell; this metadata was not recorded at the time. A third swabbing event was not included to compare the antigen test per the manufacturers’ instructions to minimize participant discomfort and as the primary focus was usability and not specific assay performance. Instead, further evaluation of the effect of the SafeSwab on antigen test results was performed by diluting NP swab fluid. Two samples from patients with acute illness (Ct 13.77 and 18.44, Alinity m SARS-CoV-2 assay) were diluted in a 1:10 series and then 10 μL or 30 μL was absorbed on to the SafeSwab and used to run an antigen test. The Ct 13.77 sample was positive neat and at dilutions 1:10 and 1:100 and the Ct 18.44 sample was positive neat and weakly positive at dilution 1:10 (Figure 3). A clear positive result was therefore obtained here when using 0.3% of a Ct 13.44 NP sample and 1% of a CT 18.44 sample.

**Figure 3.**
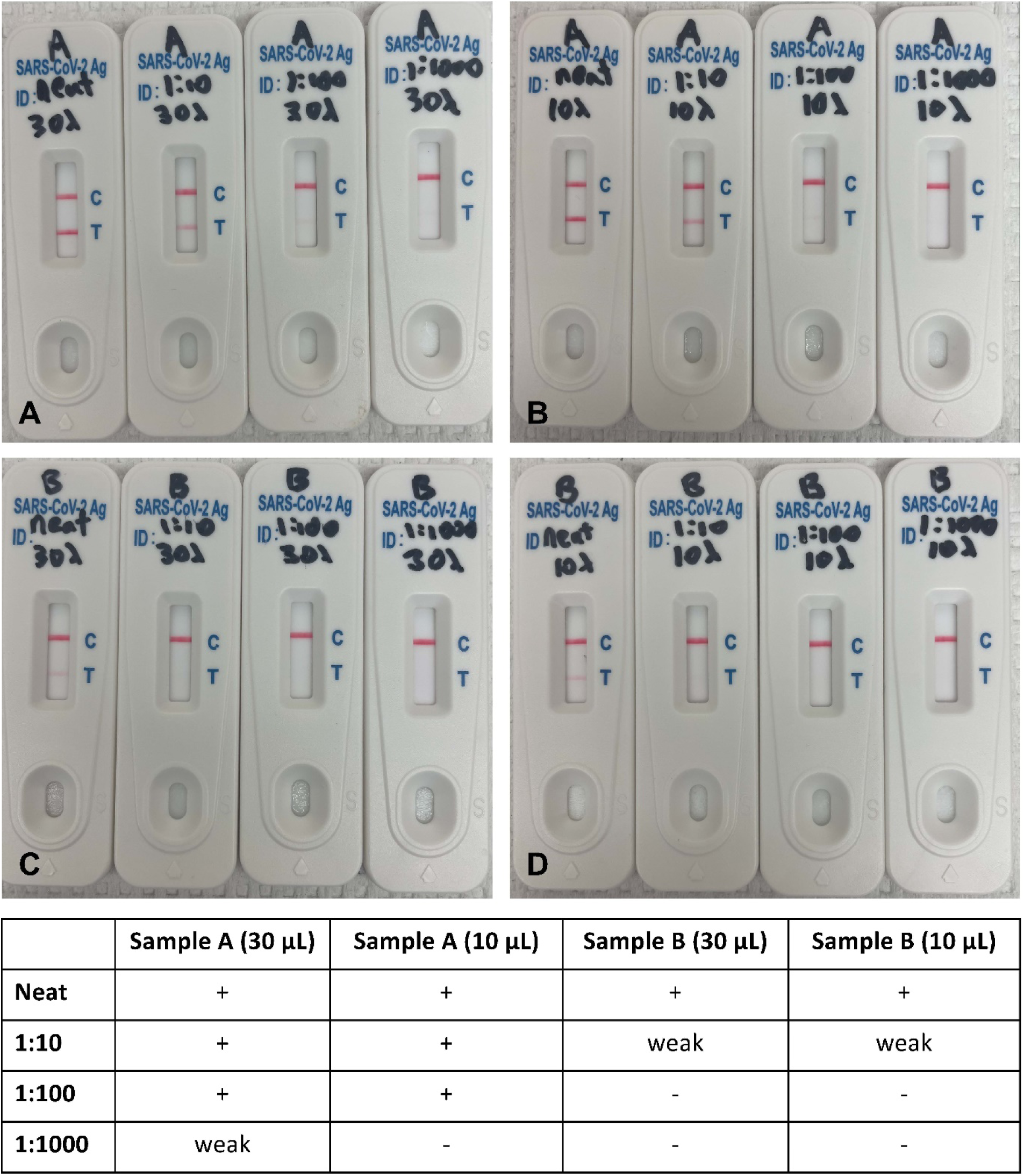
Tenfold dilution series of fluid from two nasopharyngeal swab samples with Ct 13.77 (A and B) and 18.44 (C and D); tested by absorbing 30 μL (A and C) and 10 μL (B and D) of each dilution onto the tip of the SafeSwab.

All 32 participants in the antibody testing arm produced a valid result (control line present) and of these, 32 were positive for IgG while 3 were negative. The AutoAdapt LFA algorithm correctly interpreted all 32 results compared to a human observer, including the three IgG-negative samples. Patients participated 6-10 weeks after their onset of symptoms. For this reason, despite the inclusion of separate IgM and IgG zones on the ACON SARS-CoV-2 IgG/IgM Rapid Test cartridge, we only evaluated IgG results, as IgM is known to be sporadically detected more than 4 weeks following infection (8). Venous blood was not collected to compare the results to an EUA approved antibody assay to minimize participant discomfort and as the primary focus was usability rather than specific assay performance.

The usability survey was completed by 21 (53%) antigen testing participants and 25 (78%) antibody testing participants (Table 1). Although some (24-28%) of respondants felt that they would like help collecting the sample, 100% felt that the instructions for transfering the sample from the SafeSwab into the cartridge were clear, and ultimately, 96% of respondants felt that they had collected a good sample, including all of the antibody testing. Surprisingly, those who thought that they would like help collecting the sample were mostly (5 of 7 for antibody and 3 of 5 for antigen) from the group who self-identified as having medical or laboratory training. This may reflect a self-awareness about the importance of sample collection.

**Table 1.** Patient survey results after having used the SafeSwab and HealthCheck app. Note that 21 (53%) antigen testing participants and 25 (78%) antibody testing participants opted to complete the survey and one antibody testing participant did not answer two of the questions.

| Questions on Usability | Antigen<br>Yes (%) |  | Antibody<br>Yes (%) |  | Total<br>Yes (%) |
| --- | --- | --- | --- | --- | --- |
| *Do you feel confident that you collected the sample properly?* | 20/21 | (91%) | 25/25 | (100%) | 96% |
| *Do you have any laboratory or medical training?* | 9/21 | (43%) | 7/24 | (29%) | 36% |
| *Did you feel that you needed help while collecting your sample?* | 5/21 | (24%) | 7/25 | (28%) | 26% |
| *Were instructions clearly explained how to collect the sample?* | 17/21 | (81%) | 24/24 | (100%) | 91% |
| *Were instructions clearly explained how to transfer the sample to the kit?* | 21/21 | (100%) | 25/25 | (100%) | 100% |
| *I understood how to use the lancet* | N/A |  | 25/25 | (100%) | 100% |
| *I was able to collect enough blood to make the end of the swab red* | N/A |  | 25/25 | (100%) | 100% |
| Educational Level | Number (%) |  | Number (%) |  | Total (%) |
| Doctorate | 1 | (5%) | 0 | (0%) | 2% |
| MS | 1 | (5%) | 5 | (20%) | 13% |
| BS/BN | 16 | (76%) | 14 | (56%) | 65% |
| High School or below | 3 | (14%) | 6 | (24%) | 22% |
| Age | Number (%) |  | Number (%) |  | Total (%) |
| 18-29 | 4 | (19%) | 0 | (0%) | 9% |
| 30-39 | 4 | (19%) | 4 | (16%) | 17% |
| 40-49 | 1 | (5%) | 4 | (16%) | 11% |
| 50-59 | 6 | (29%) | 5 | (20%) | 24% |
| 60-69 | 3 | (14%) | 9 | (36%) | 26% |
| 70-79 | 3 | (14%) | 3 | (12%) | 13% |

Here we introduce SafeSwab, a new testing device that enables sampling of an anterior nasal sample or a fingerstick blood sample, and with an integrated buffer chamber, enables easy delivery of the sample and buffer into a testing device. The SafeSwab is ideal for near-patient testing such as with a point-of-care device. We implemented the SafeSwab in conjunction with the Safe Health Systems HealthCheck digital health application to determine if patients could perform an antigen test after taking their own anterior nasal sample, or a serology test after collecting their own fingerstick blood. In a typical antigen test, the swab is placed into a buffer tube, swirled for a period of time, then removed, and a dropper cap placed on top. Then drops are placed into the test cartridge. With the SafeSwab, these multiple steps are integrated into one device. In an antibody test, the blood is typically transferred to the sample well of the cartridge with a miniature plastic pipette or capillary tube, both of which require skill and dexterity. Then, buffer must be measured into the sample well. We see the primary advantages of the SafeSwab to be reduced chance of user error, such that it is easy enough for a lay user.

Despite all patients having documented PCR results, three antibody tests were negative. A previous study showed this antibody test to be highly accurate in a set of samples drawn in the first 1-4 weeks after diagnosis (9). It is possible that antibody levels in these three patients had decreased, or that they had a low response; variability in anti-SARS-CoV-2 IgG response is well documented (10). Combining valid results of both antigen and antibody tests, 63 of 69 (91%) patients generated the correct result compared to the EUA approved PCR reference standard.

From an operational standpoint, 69 of 72 (96%) patients generated a valid result. In all cases (n=72), the AutoAdapt LFA algorithm interpreted the test correctly when compared to the human interpretation. Design and performance characteristics of this algorithm are outlined in a recent preprint (2).

In this study, the Safe Health System HealthCheck application was used to guide the participant through the testing steps and transfer the images to the AudoAdapt LFA algorithm. This digital health platform has the capacity to integrate features such as provision of reliable medical information, symptom checking algorithms, connection to healthcare providers through virtual consults, self-testing instructions, automated test interpretation (using the AutoAdapt LFA program), integration of test results into a patient record (if the patient chooses to link accounts), and public health reporting for reportable diseases. These features, when paired with the SafeSwab, could enable and accelerate new models of patient care, including increasing access to testing and decreasing cost of healthcare.

## Data Availability

The data generated in this study is available in this publication. Additional data availability is not applicable in this case

## Acknowledgements

We would like to thank the hard work of the research coordinators and study nurses who supported the study. We also acknowledge ACON Laboratories (San Diego, CA) for supplying kits for evaluation.

## Notes

### Competing Interest Statement

TEG represents Mayo Clinic in a joint venture with Safe Health Systems and has shared intellectual property that may result in royalty sharing. Safe Health Systems has licensed the AutoAdapt LFA scanning algorithm from Columbia University. This work was partially a result of sponsored research funded by Safe Health Systems.

### Funding Statement

This study was supported by the Mayo Clinic Center for Individualized Medicine and funds from the Geraldine Colby Zeiler Professorship funds. The work was supported by Herbert Irving Comprehensive Cancer Center in partnership with the Irving Institute for Clinical and Translational Research via a SARS-CoV-2 Research Pilot Grant, Columbia University School of Engineering and Applied Science via a Technology Innovations for Urban Living in the Face of COVID-19 Pilot Grant, and a gift from Bing Zhao to Columbia University.

### Author Declarations

The Mayo Clinic Institutional Review Board reviewed and approved protocols 20-004544 and 20-010688 for informed consent of participants into this study.

