## Supplemental Figure 1 for "SafeSwab, a sample collection and dispensing device for near-patient testing"

| | Valid True Positive | Valid True Negative | Valid False Positive | Valid False Negative | Invalid |
|---|---|---|---|---|---|
| Flowflex™ SARS-CoV-2 Antigen Rapid Test | 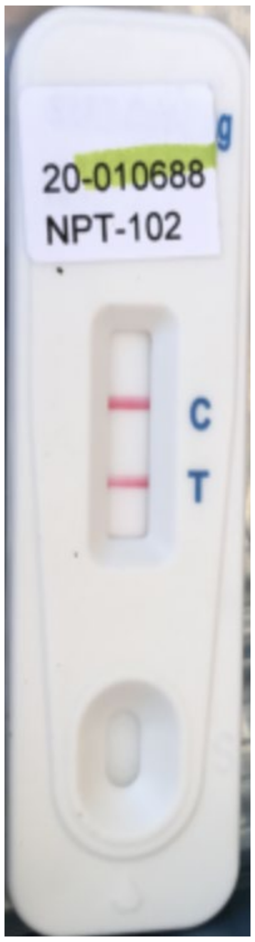 | 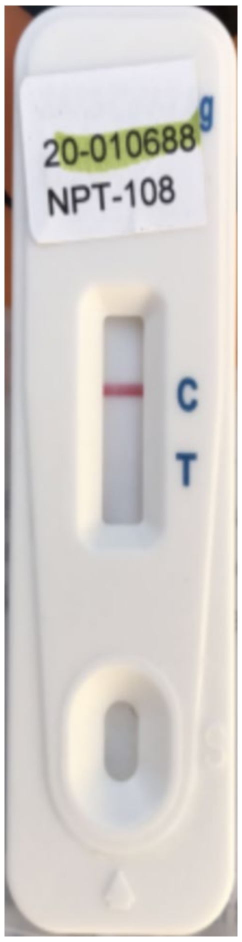 | 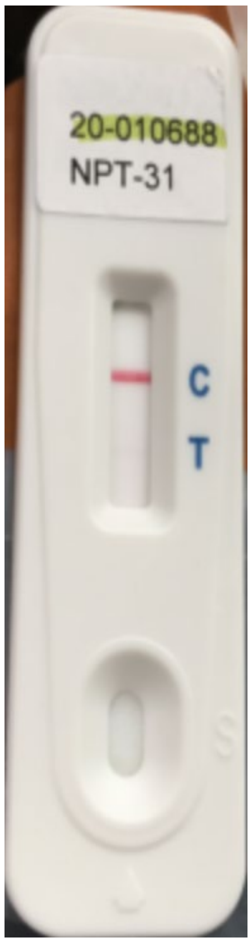 | 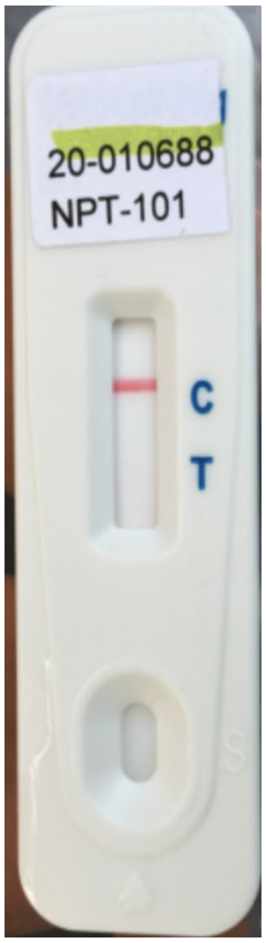 | 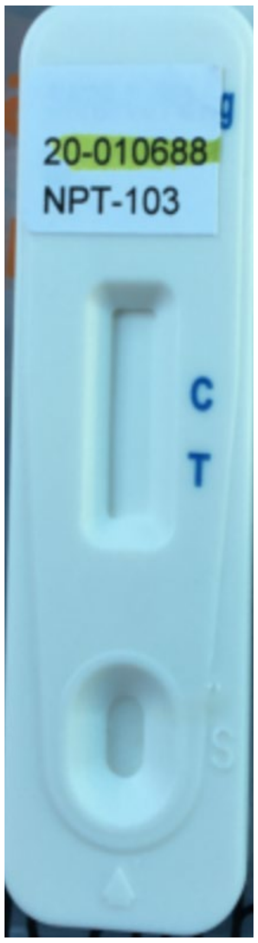 |
| ACON SARS-CoV-2 IgG/IgM Rapid Test | 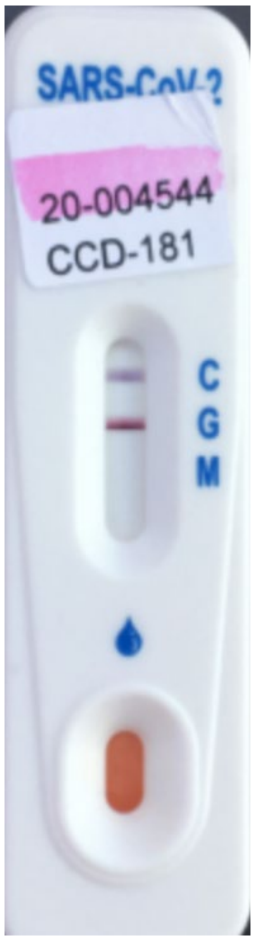 | N/A | N/A | 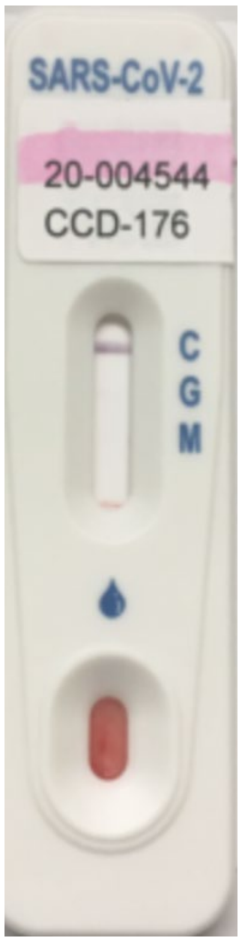 | N/A |

**Supplementary Figure 1.** Representative images from each recorded test outcome. Note that the false positive antigen result is difficult to see in this unenhanced image; however, both the manual interpreter and the AutoAdapt algorithm independently interpreted this test as positive.
